# DEVELOPING NEW DUAL-ACTION ANTIVIRAL/ANTI-INFLAMMATORY SMALL MOLECULES FOR COVID-19 TREATMENT USING IN SILICO AND IN-VITRO APPROACHES

**DOI:** 10.1101/2024.11.06.24316825

**Authors:** Vladimir V. Ivanov, Anton B. Zakharov, Dmytro O. Anokhin, Olha O. Mykhailenko, Sergiy M. Kovalenko, Larysa V. Yevsieieva, Victoriya A. Georgiyants, Michal Korinek, Yu-Li Chen, Shu-Yen Fang, Mohamed El-Shazly, Tsong-Long Hwang, Oleg M. Kalugin

## Abstract

This study aims to develop new molecular structures as potential therapeutic agents against COVID-19, utilizing both *in silico* and *in vitro* studies. Potential targets of cepharanthine (CEP) against COVID-19 to reveal its underlying mechanism of action were evaluated using *in silico* screening experiments. A library of new molecules was docked into the receptor binding domain of the SARS-CoV-2 spike glycoprotein complex with its receptor, human ACE2, to identify promising compounds. Receptor-oriented docking was performed using the most likely macromolecular targets, aimed at inhibiting key viral replication pathways and reducing inflammatory processes in damaged tissues. The hit molecules showed potential inhibition of Mpro and PLpro proteases of SARS-CoV-2, which are involved in viral replication. They also showed a potential inhibitory effect on Janus kinase (Jak3), which mediates intracellular signaling responsible for inflammatory processes.

The *in vitro* study examined the effects of the selected hit molecules on the generation of superoxide anions and the release of elastase in activated neutrophils, which are factors that exacerbate tissue inflammation and worsen the clinical manifestations of COVID-19. It was demonstrated that 2-((5-((4-isopropylphenyl)sulfonyl)-6-oxo-1,6-dihydropyrimidin-2-yl)thio)-N-(3-methoxyphenyl)acetamide (**Hit15**) inhibited virus infection by 43.0% at 10 *μ*M using pseudovirus assay and suppressed fMLF/CB-induced superoxide anion generation and elastase release in human neutrophils with IC_50_ values 1.43 and 1.28 *μ*M, respectively. **Hit15** showed promising activity against coronavirus that can be further developed into a therapeutic agent.

**Graphical Abstract:** 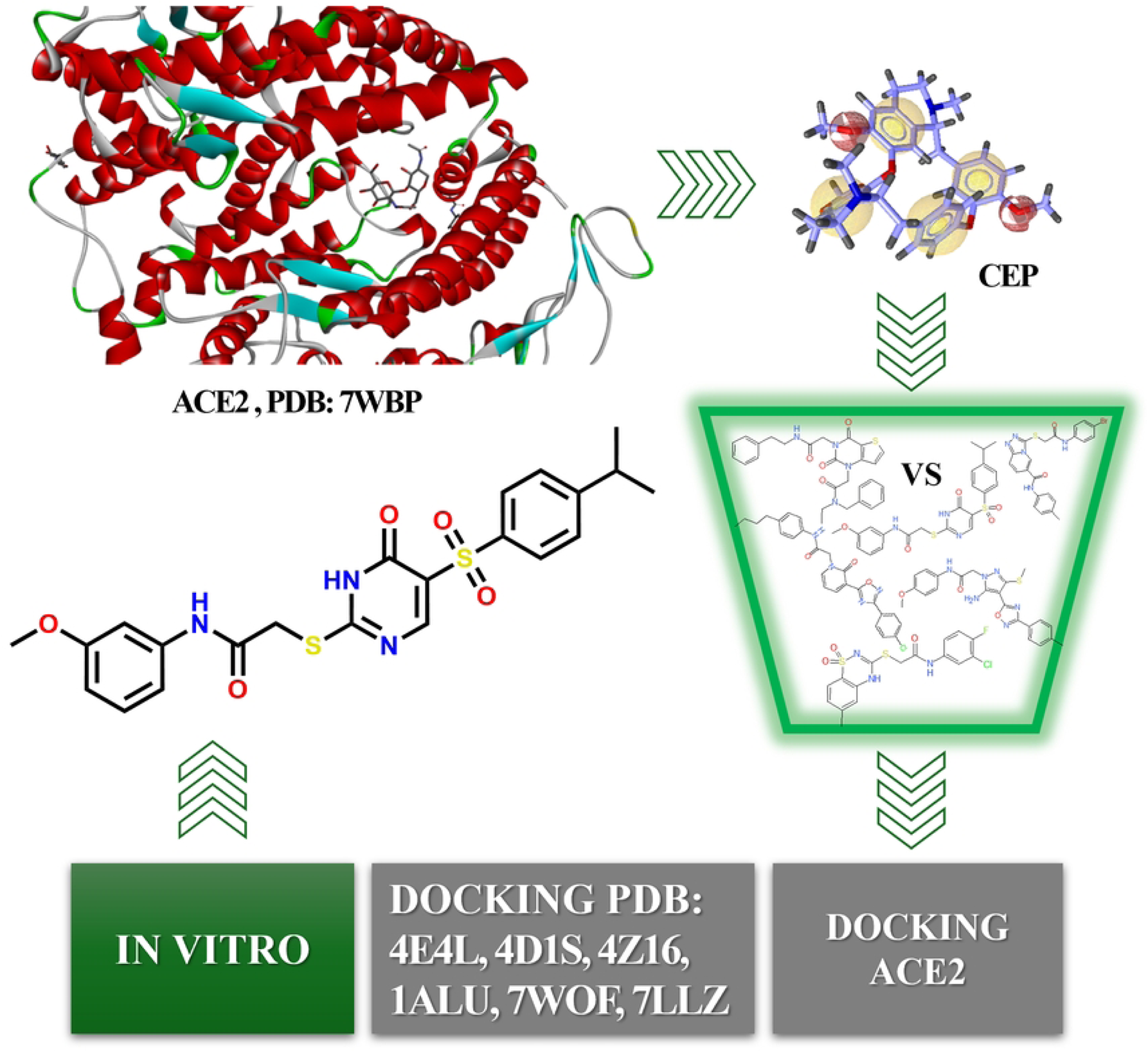

## Introduction

The recent trends in the search for effective drugs against COVID-19 focus on the development of inhibitors of key SARS-CoV-2 proteins. Integrating antiviral and anti-inflammatory therapy is a critical approach to managing COVID-19 since the inflammation caused by the immune response leads to serious complications of the disease [1, 2].

Understanding the pathological progression and clinical manifestations of COVID-19 is a prerequisite for the development of drugs for rational therapeutic intervention. The challenge of developing effective drugs to combat SARS-CoV-2 remains unresolved. There is an urgent need for medications that can suppress the main mechanisms of SARS-CoV-2 replication within the cell and mitigate the consequences of its impact on the human body [3]. This need has become especially relevant with the emergence of new viral variants capable of evading immune protection provided by vaccines [4, 5].

COVID-19, caused by the SARS-CoV-2 virus, is a complex, multi-organ, and heterogeneous disease. The clinical manifestations of COVID-19 are diverse. The disease can progress from uncomplicated forms to pneumonia and acute respiratory distress syndrome (ARDS), requiring intensive care [6]. The pathogenesis of COVID-19 is driven by a hyperactive inflammatory response, leading to severe inflammation and tissue damage, particularly in the lungs [7]. It is known that excessive neutrophil activation causes tissue damage, and the neutrophil activation pathway plays a key role in the poor prognosis of COVID-19 patients by exacerbating lung inflammation and respiratory failure. Activated neutrophils secrete several cytokines, including superoxide anion, which can directly or indirectly cause tissue damage. Neutrophil elastase is a major product secreted by activated neutrophils and a key factor in tissue destruction in inflammatory diseases [8]. The inhibition of superoxide formation and elastase release, which are the markers of the anti-inflammatory response, can reduce the inflammatory burden and limit damage to the lungs and other tissues caused by COVID-19 [9].

The integration of antiviral and anti-inflammatory therapies is a key approach to managing COVID-19. This approach was initially utilized, for example, with the introduction of corticosteroids into treatment regimens to reduce the body’s inflammatory response [1, 2]. In this context, there is growing interest in biologically active substances from the plant *Jordanian hawksbeard*, as they possess both anti-inflammatory and antiviral properties. These substances inhibited the main protease of SARS-CoV-2 and reduced inflammatory processes [8, 10].

Studies confirmed the *in vitro* and *in vivo* antiviral potential of cepharanthine (CEP) against SARS-CoV-2. CEP demonstrated multiple molecular mechanisms, including the suppression of the viral entry phase (by interacting with the viral spike protein and blocking its binding to ACE2) and reducing the production of inflammatory factors [11, 12].

Approaches that enable the targeted creation of new pharmacologically active molecules were implemented in previous studies including computer-aided molecular modeling (CAMM) and QSAR methods [13]. In our search for new agents against SARS-CoV-2, we used workflows that combine several tools, such as pharmacophore screening of large chemical spaces and molecular docking of preselected candidates [14].

In this study, we used *in silico* screening experiments and receptor-oriented docking in the receptor binding domain of the SARS-CoV-2 Omicron spike glycoprotein complex with its receptor, human ACE2. We also used the three-dimensional structural models of other active sites of biological molecules involved in the mechanisms of SARS-CoV-2’s impact on the body, using cepharanthine as a representative structure.

## Materials and Methods

### Computational Procedures

Molecular ligand analysis and the design of new biologically active molecules were performed using free and publicly available software packages including Jmol [15], PyMol [16], and LigandScout 4.4 [17, 18], which allowed for the pharmacophore analysis and virtual screening of specific molecular databases relative to the generated pharmacophore.

LigandScout tools [18] were used to identify the molecular parameters that would correspond to drug-like properties. Molecules with poor permeability and oral absorption (molecular weight > 500, C logP > 5, more than five hydrogen bond donors, and more than ten acceptor groups) were excluded [19].

To understand the possible mechanisms of interaction between substances and biological targets, we used the SwissTargetPrediction web server [20] to predict the biological activity of small molecules. The DataWarrior program was employed to calculate physicochemical properties and analyze molecular scaffolds [21].

### Preparation of Proteins and Database

X-ray crystal structures of the target proteins were obtained from the Protein Data Bank (PDB) [22] and used for virtual screening and receptor-oriented docking. Molecules with properties matching the pharmacophore structure constituting a set of promising molecules ("hits"), were used in the docking procedure with the active site of the corresponding target protein.

For the *in silico* virtual screening, we used a chemical space of molecules containing more than 70,000 organic compounds. The compounds of this database (DB_KSM) were synthesized by the synthetic group of Prof. S.M. Kovalenko (Department of Organic Chemistry at Kharkiv National University, Ukraine). They included various heterocyclic systems with substituents of different electronic nature, including various aliphatic fragments, amino acid residues, halogens, and others. The representative structure was cepharanthine (CEP), CAS Number 481-49-2 [23].

### Molecular Docking

X-ray crystal structures of the corresponding proteins from the Protein Data Bank were used for docking. Receptor-oriented docking was performed using the AutoDock Vina program [24].

### Chemistry

The synthesis of potentially active hit compounds from virtual screening and docking experiments was carried out using previously developed methods. **Hit2** [25], **Hit3** та **Hit5** [26], **Hit9**, **Hit10** [27], **Hit13** [28], **Hit7**, **Hit15** [29] were prepared (**Fig. 1**).

**Fig 1.**
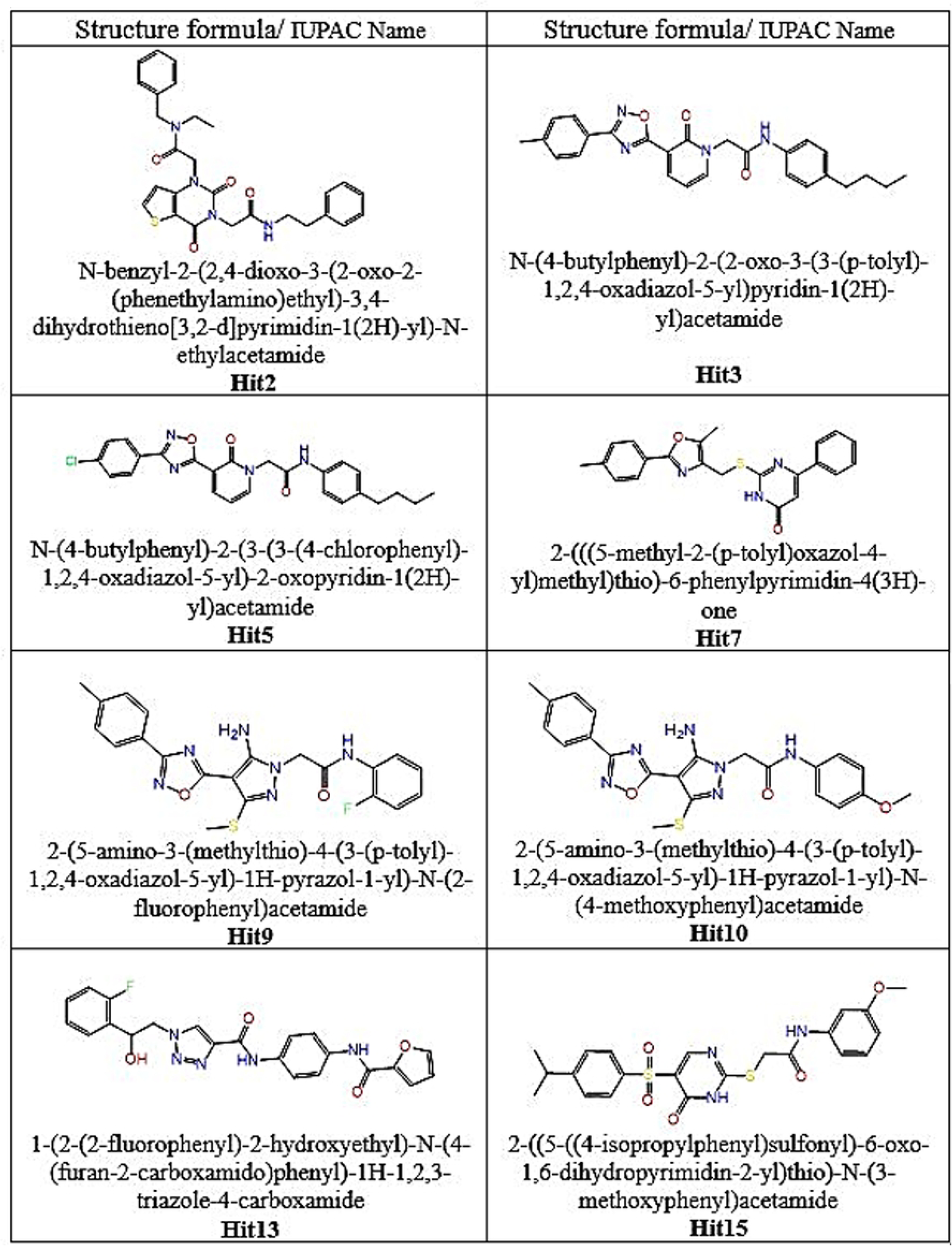
Potentially active hits selected as a result of virtual screening and docking. All NMR spectra were recorded on a Varian MR-400 spectrometer with standard pulse sequences operating at 400 MHz for ^1^H NMR and 101 MHz for ^13^C NMR. For all NMR spectra, DMSO-d*_6_* was used as the solvent. Chemical shift values are referenced to residual protons (*δ*_H_ 2.49 ppm) and carbons (*δ*_C_ 39.6 ppm) of the solvent as an internal standard. LC/MS spectra were recorded on an ELSD Alltech 3300 liquid chromatograph equipped with a UV detector (*λ*_max_ 254 nm), API-150EX mass-spectrometer, and using a Zorbax SB-C18 column, Phenomenex (100 × 4 mm) Rapid Resolution HT cartridge 4.6 × 30 mm, 1.8-Micron. Elution started with 0.1 M solution of HCOOH in water and ended with 0.1 M solution of HCOOH in acetonitrile, using a linear gradient at a flow rate of 0.15 mL/min and an analysis cycle time of 25 min.

### *In vitro* anti-inflammatory activity in human neutrophils

For the *in vitro* studies, the synthesized compounds (**Fig. 1**) were dissolved in dimethyl sulfoxide (DMSO) to prepare stock solutions. The final concentration of DMSO in cell experiments did not exceed 0.5% and did not affect the measured parameters. Blood was taken from healthy human donors using a protocol approved by the Chang Gung Memorial Hospital review board. Neutrophils were isolated following the standard procedure [30].

The inhibition of superoxide anion generation (respiratory burst) was measured based on ferricytochrome *c* reduction as previously described [31]. Briefly, preheated neutrophils (6 × 10^5^ cells·mL^−1^) and 0.6 mg/mL ferricytochrome *c* solution were treated with the tested compounds or DMSO (control) for 5 min. The cells were activated with formyl-methionyl-leucyl-phenylalanine (fMLF, 100 nM)/cytochalasin B (CB, 1 μg/mL) for 13 min. The absorbance was continuously monitored at 550 nm using Hitachi U-3010 spectrophotometer with constant stirring (Hitachi Inc., Tokyo, Japan). Calculations were based on the differences in absorbance with and without superoxide dismutase (SOD, 100 U/mL) divided by the extinction coefficient for the reduction of ferricytochrome *c* (ε = 21.1/mM/10 mm).

Elastase release (i.e., degranulation from azurophilic granules) was evaluated as described before [32]. Briefly, neutrophils were equilibrated with elastase substrate, MeO-Suc-Ala-Ala-Pro-Val-*p*-nitroanilide (100 μM), at 37 °C for 2 min and then incubated with the sample for 5 min. Cells were activated by 100 nM fMLF and 0.5 μg/mL CB for 13 min, and changes in the absorbance at 405 nm corresponding to elastase release were continuously monitored. The results were expressed as the percentage of the initial rate of elastase release in the fMLF/CB-activated drug-free control system.

### Pseudotyped lentivirus assay

Lentivirus experiments were approved by the Institutional Biosafety Committee of Chang Gung University and performed according to our previous report [42]. Stable hACE-2 overexpressed HEK293T cells were provided by Dr. Rei-Lin Kuo (Chang Gung University) and maintained in DMEM containing 10 % FBS and 10 µg/mL blasticidin. VSV-G pseudotyped lentivirus control (clone name: S3w.Fluc.Ppuro) and SARS-CoV-2 S-protein expressing VSV-G pseudotyped lentiviruses (clone name: nCoV-S-Luc-D614G and nCoV-S-Luc-B.1.617.2) were purchased from RNAi Core Facility of Academia Sinica. hACE-2 overexpressed cells (1 × 10^4^ cells/well) were seeded on 96-well plates and incubated at 37 ℃ with 5% CO_2_. Equal relative infection units (RIU) (5 × 10^3^ RIU/well= 0.5 RIU/cell) of pseudotyped lentiviruses were pretreated with various concentrations of antiviral agents in DMEM containing 5% FBS at 37 ℃ for 1 hour. The medium of ACE-2 overexpressed cells was replaced with treated pseudotyped lentivirus and cultured for 24 h. Luciferase activity was measured using a Luciferase Assay System (E2520, Promega) and recorded by a Fluorescence Reader. Cepharanthine served as a positive control.

### WST-1 viability assay

The potential cytotoxicity of the tested samples was evaluated by the WST-1 reduction assay in hACE-2-overexpressed HEK293T [40]. The hACE-2-overexpressed HEK293T cells (1 × 10^4^ cells/well) were preincubated with DMSO or tested agents for 24 h. Then, the WST-1 reagent (M192427, Sigma-Aldrich, MO, USA) was added and incubated for 4 h at 37 °C. The absorbance at 405 nm was measured by a Multiska GO spectrophotometer (Thermo Fisher Scientific, MA, USA).

### Statistical analysis

Results are expressed as mean ± S.E.M. of two independent measurements (pseudoviral anti-viral assay) or mean ± S.E.M. of three independent measurements (anti-inflammatory assay). The 50% inhibitory concentration (IC_50_) was calculated from the dose-response curve obtained by plotting the percentage of inhibition versus concentrations (linear function, Microsoft Office, anti-inflammatory assay). Statistical analysis was performed using Student’s *t*-test (Sigma Plot, Systat Software, Systat Software Inc., anti-inflammatory). Values with \**P* < 0.05, \*\**P* < 0.01, \*\*\**P* < 0.001 were considered statistically significant.

## Results

### *In silico* models and calculations

Studies of the bis-benzylisoquinoline alkaloid cepharanthine (CEP) against COVID-19 identified its significant antiviral and anti-inflammatory potential. However, the precise mechanism of CEP is still under investigation [11, 12]. To model the mechanism of cepharanthine’s action on COVID-19, researchers focused on various protein targets available in the Protein Data Bank (PDB). According to scientific literature, one of the primary mechanisms of CEP’s action against COVID-19 is its binding to ACE2, which was validated by molecular dynamics (MD) simulations [33]. Cepharanthine was selected as the representative structure to develop new molecules with the potential to inhibit key viral pathways. The 3D crystal structure of the receptor binding domain of the SARS-CoV-2 Omicron variant spike glycoprotein complex with its receptor, human ACE2 (PDB: 7WBP), was chosen as the biological target.

We conducted a preliminary pharmacophore screening using the LigandScout suite to identify suitable structures for further investigation. The receptor-binding domain of the SARS-CoV-2 Omicron spike glycoprotein complex with the human ACE2 receptor and the ligand NAG PDB 7WBP [34] was employed as a representative protein-ligand structure for the study (**Fig. 2**).

**Fig. 2.**
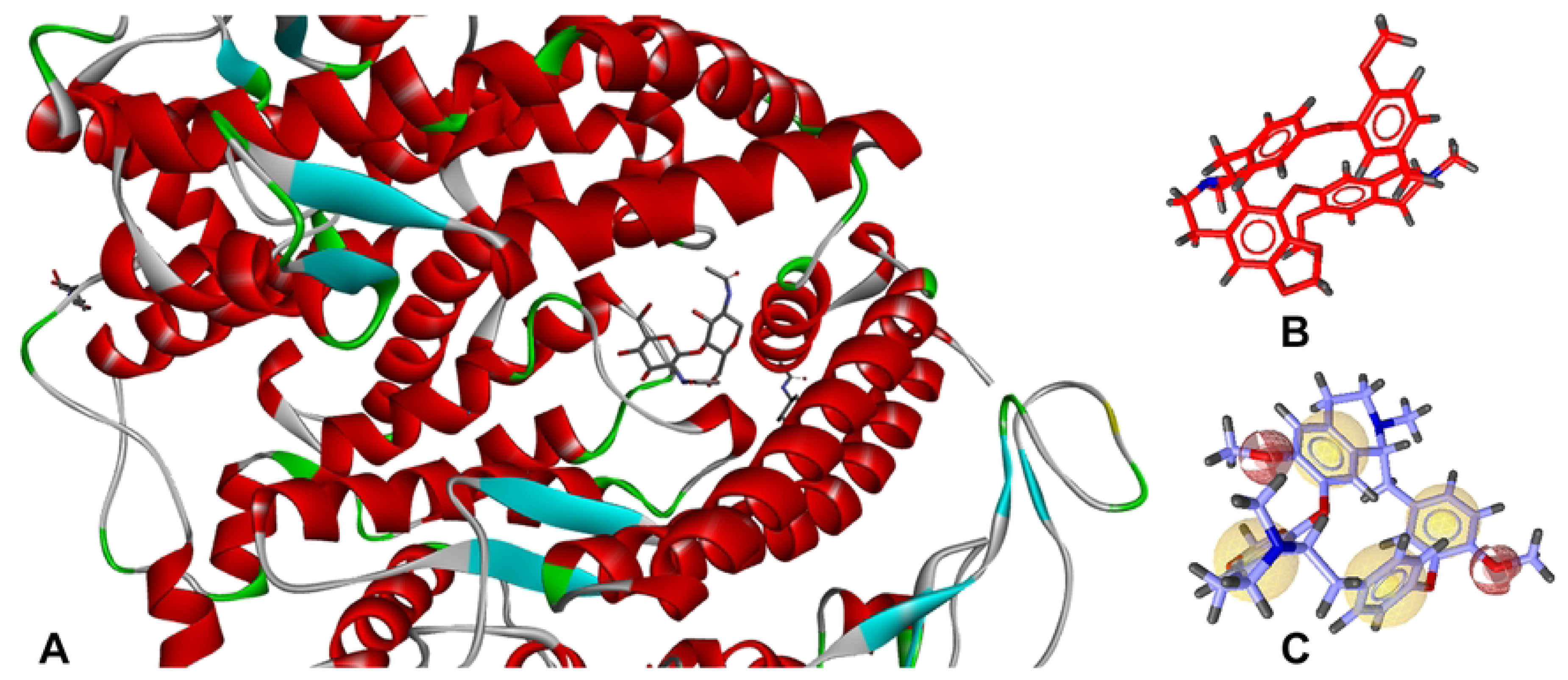
Receptor binding domain of SARS-CoV-2 Omicron spike glycoprotein complex with its receptor human ACE2 with ligand NAG (A), the structure of cepharanthine CEP (D) and its pharmacophore (C).

The pharmacophore model CEP was used as a search query to identify compounds targeting the binding site of the biological target. Virtual screening (VS) of a molecular database (DB_KSM) according to the pharmacophore identified in the previous step resulted in identifying several compounds that could be promising in the fight against COVID-19. Based on matching with the pharmacophore, 23 compounds were selected for further study (**Fig. 3**).

**Fig 3.**
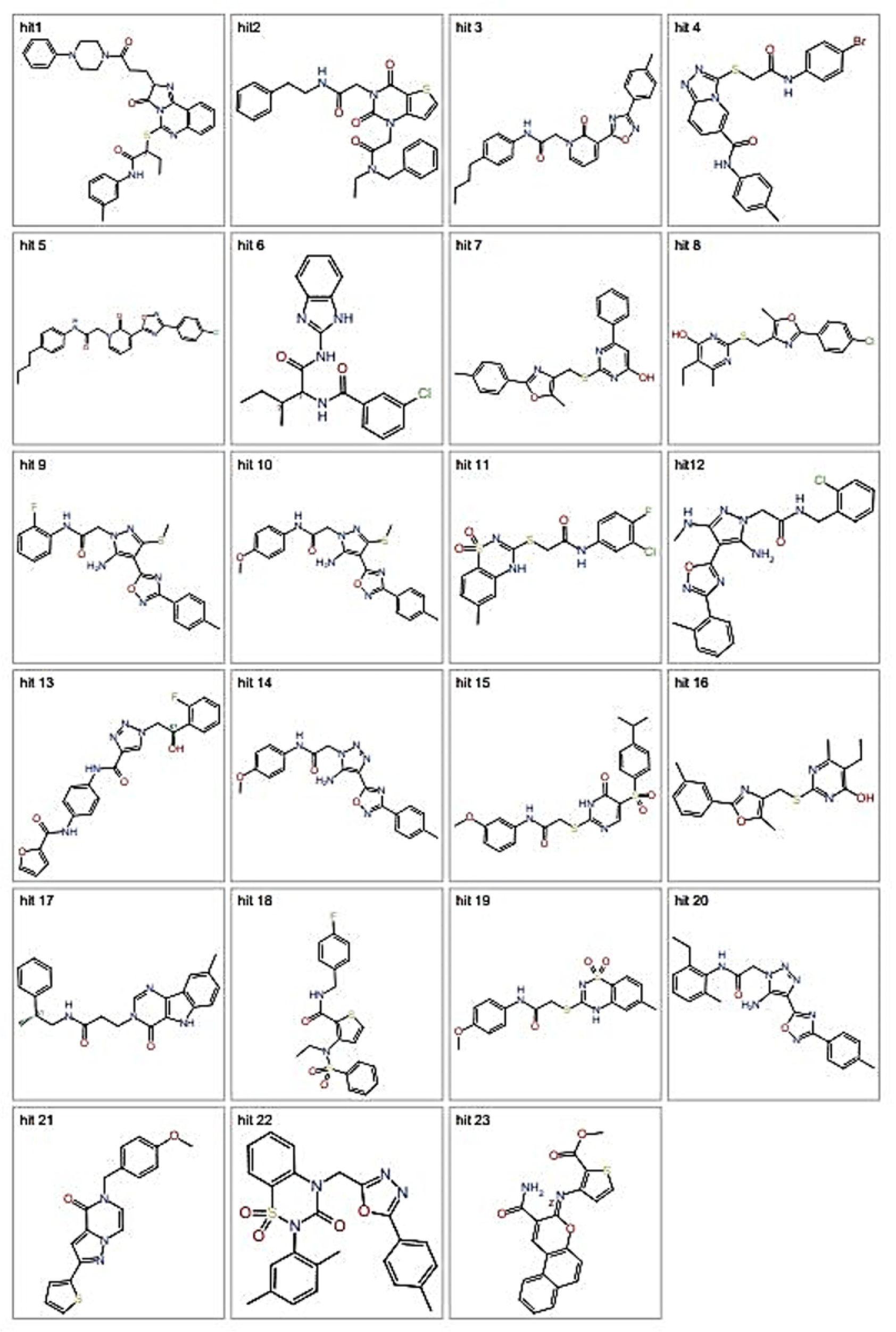
The structures with possible activity against SARS-COV2 according to VS results of DB_KSM database.

The docking of these molecules to the receptor-binding domain of the SARS-CoV-2 Omicron spike glycoprotein complex with the human ACE2 receptor (PDB 7WBP) was performed. Their binding affinity parameters were calculated. The binding affinity scores (BAS) and binding energies (BE, kcal/mol) of the docked hit compounds, along with the co-crystallized ligand (NAG) and CEP, are presented in **Table 1**.

**Table 1.** Affinity parameters of hit structures (complex PDB 7WBP)

| Mol | BAS | BE<br>(kcal/mol) | Mol | BAS | BE<br>(kcal/mol) |
| --- | --- | --- | --- | --- | --- |
| NAG | -2.04 | -7.10 | CEP | -17,54 | -7,30 |
| **Hit1** | -6,93 | -6,60 | **Hit13** | **-17.71** | **-9.10** |
| **Hit2** | **-11.14** | **-8.50** | **Hit14** | -6,88 | -6,50 |
| **Hit3** | **-15.91** | **-7.90** | **Hit15** | **-10.14** | **-9.10** |
| **Hit4** | -6,78 | -7,10 | **Hit16** | -6,56 | -7,10 |
| **Hit5** | **-12.41** | **-7.60** | **Hit17** | -6,38 | -6,90 |
| **Hit6** | -5,86 | -7,00 | **Hit18** | -5,85 | -6,70 |
| **Hit7** | **-7.56** | **-8.60** | **Hit19** | -5,49 | -6,80 |
| **Hit8** | -7,00 | -6,90 | **Hit20** | -7,13 | -7,00 |
| **Hit9** | **-10.78** | **-8.50** | **Hit21** | -6,67 | -6,56 |
| **Hit10** | **-13.89** | **-8.40** | **Hit22** | -2,54 | -6,10 |
| **Hit11** | -6,48 | -6,90 | **Hit23** | -4,34 | -5,85 |
| **Hit12** | -5,26 | -7,00 |  |  |  |

As shown in **Table 1**, eight compounds (**Hit2**, **Hit3**, **Hit5**, **Hit7**, **Hit9**, **Hit10**, **Hit13**, and **Hit15**) exhibited binding energies exceeding the absolute values of the representative cepharanthine (CEP) structure. These compounds were selected for further molecular docking studies with target proteins identified in the literature as potential pharmacological targets of cepharanthine [33].

For the calculations, hub targets for the potential pharmacological mechanism of CEP against COVID-19 were selected (**Table 2**). In the study [33], "network pharmacology" combined with RNA sequencing, molecular docking, and MD modeling was used to identify hub targets and potential pharmacological mechanisms of cepharanthine (CEP) against COVID-19. Nine key hub genes (ACE2, STAT1, SRC, PIK3R1, HIF1A, ESR1, ERBB2, CDC42, and BCL2L1) were identified. Based on these data, we performed docking studies of the home library to the hub targets from the Protein Data Bank [22] described above.

**Table 2.** Hub-targets and potential pharmacological mechanisms of action of cepharanthine (CEP) against COVID-19.

| **Target** | **Designation** | **Classification** |
| --- | --- | --- |
| **1BF5** | Tyrosine phosphorylated STAT-1/DNA complex | Gene regulation/DNA |
| **1A07** | C-SRC (SH2 domain) complexed with acemalonyl tyr-glu-(N,N-dipentyl amine) | Complex (transferase/peptide) |
| **2IUI** | Crystal structure of the PI3-kinase p85 N-terminal SH2 domain in complex with PDGFR phosphotyrosyl peptide | Transferase |
| **1H2K** | Factor Inhibiting HIF-1 alpha in complex with HIF-1 alpha fragment peptide | Transcription activator/inhibitor |
| **2AJF** | Structure of SARS coronavirus spike receptor-binding domain complexed with its receptor | Hydrolase / viral protein |
| **1A52** | Estrogen receptor alpha ligand-binding domain complexed to estradiol | Receptor |
| **2A91** | Crystal structure of ErbB2 domains 1-3 | Signaling protein, transferase, membrane protein |
| **7S0Y** | Structures of TcdB in complex with Cdc42 | Hydrolase |
| **7JGW** | Crystal structure of BCL-XL in complex with compound 1620116, crystal form 1 | Apoptosis |

Docking was performed using AutoDock Vina with CEP and the studied molecules. The results of molecular docking and binding energies of the molecules are presented in **Table 3**.

**Table 3.**
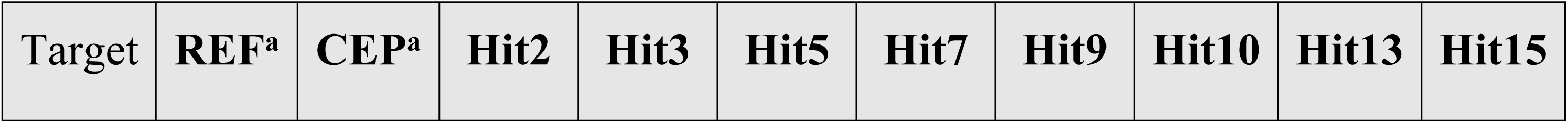
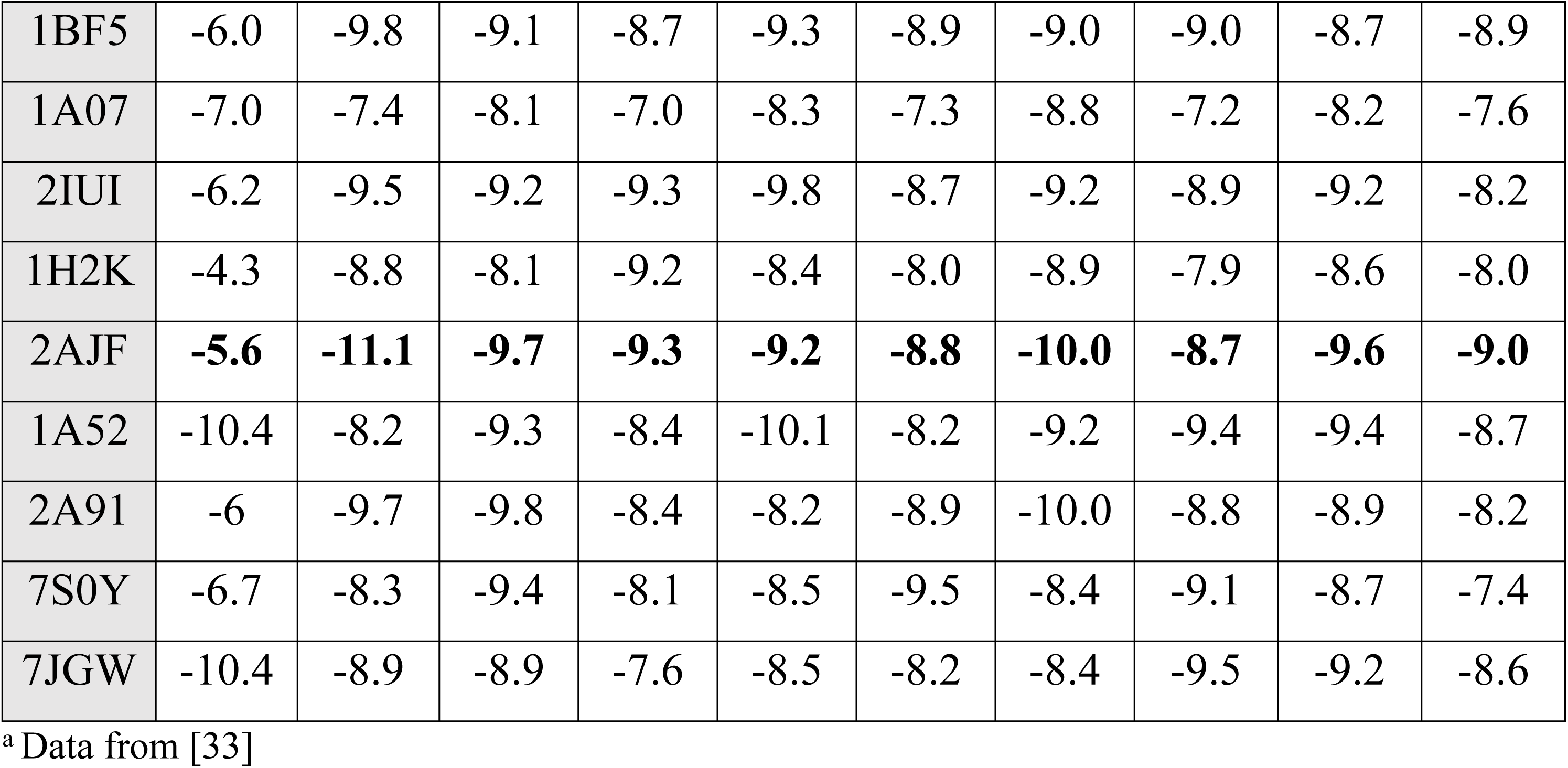
Molecular docking binding energy results of hub genes with CEP and hit-molecules.

The molecular docking results showed a good potential for inhibition of Target 2AJF, the spike receptor-binding domain of SARS-CoV-2, in complex with its receptor and a set of targets responsible for modulating the immune response.

Additionally, we utilized the SwissTargetPrediction resource further to evaluate the most likely macromolecular targets for CEP (**Fig. 4**) and the new molecules to assess the possibility of using multiple therapeutically relevant targets for the studied compounds, as most bioactive molecules have more than one target [35].

**Fig 4.**
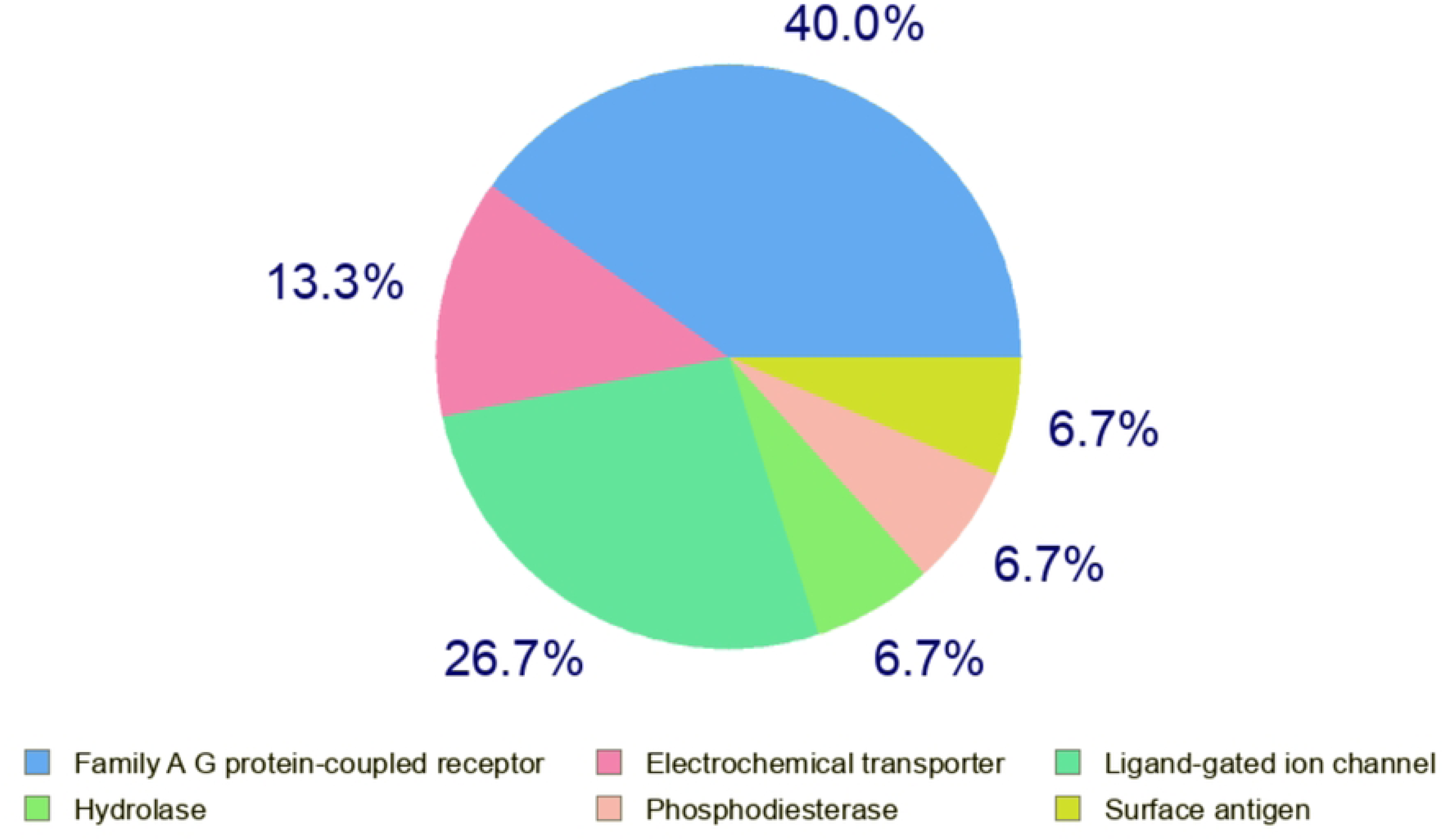
The most probable macromolecular targets of CEP, suggested by the SwissTargetPrediction resource [20].

The identified biological targets for CEP included the family A G protein-coupled receptors – cell surface proteins that detect molecules outside the cell and activate cellular responses, including immune responses. They also included the electrochemical transporters and ligand-gated ion channels responsible for signal molecule transport regulated by membrane potential or ion flow. The biological targets of the new hit compounds are presented as an aggregate bar chart (**Fig. 5**), reflecting the contribution of each category.

**Fig 5.**
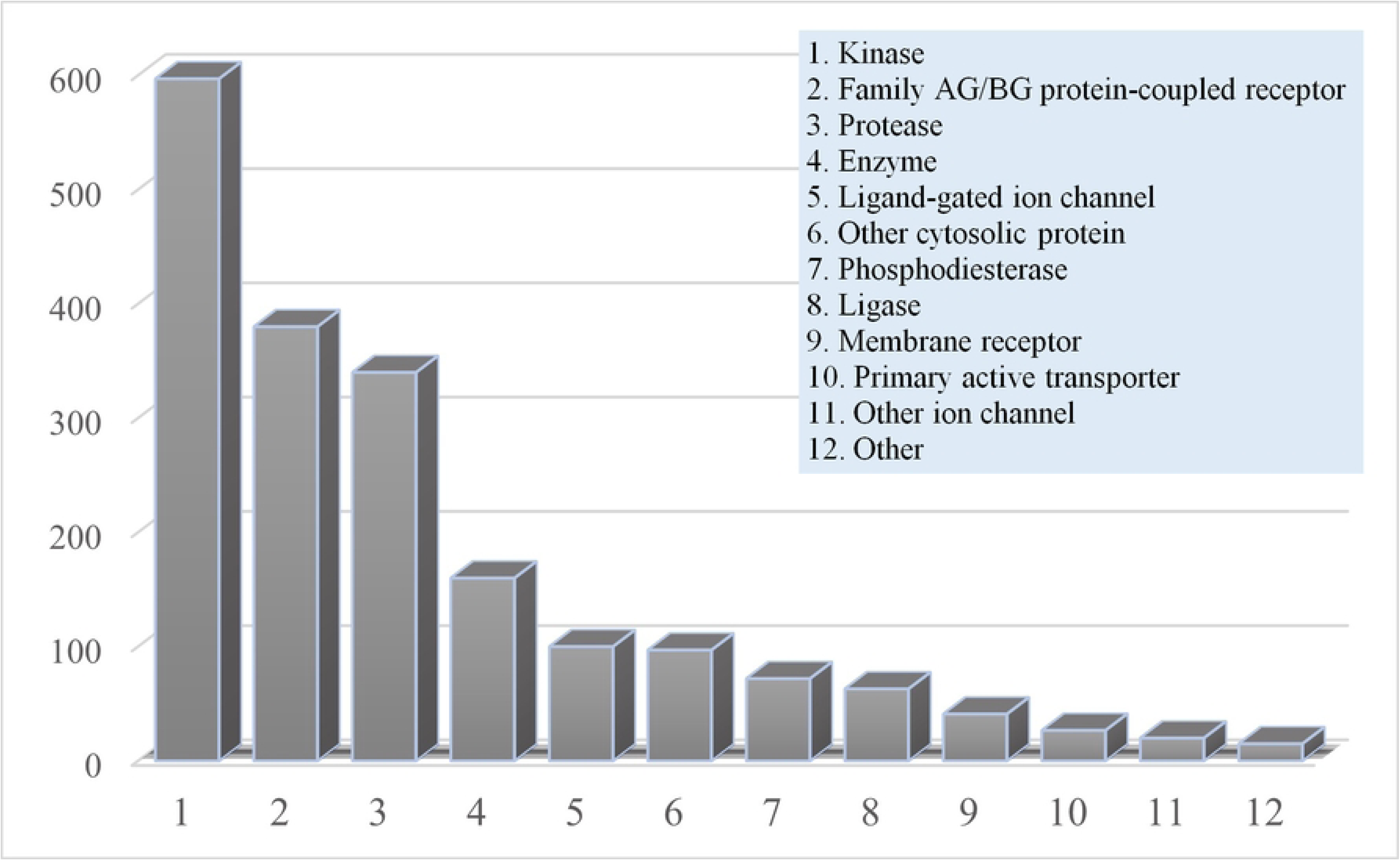
Biological target pool of the new hit molecules proposed by the SwissTargetPrediction resource [20].

The primary predicted target categories for the new hit molecules, as proposed by the SwissTargetPrediction resource, were the kinase/family A G / B G protein-coupled receptors/protease.

**Table 4** presents the categories of biological targets for the new hit molecules: a) Targets responsible for modulating the immune response, which may be one of the mechanisms contributing to tissue inflammation in COVID-19 (kinases (phosphotransferases) / protein-coupled receptor A G /B G family); b) Targets from the protease category, which may be involved in inhibiting key pathways of viral replication.

**Table 4.** Categories of biological targets for pharmacological action against SARS-COV-2 used in the study.

| Target | Designation | Classification |
| --- | --- | --- |
| 4E4L | AK1 kinase (JH1 domain) in complex | Transferase/kinase |
| 4D1S | Crystal structure of JAK2 kinase | Transferase/kinase |
| 4Z16 | Crystal Structure of the Jak3 Kinase Domain | Transferase/kinase |
| 1ALU | Human Interleukin-6 | Cytokine |
| 7WOF | Crystal structure of SARS-CoV-2 Mpro | Viral protein/protease |
| 7LLZ | Crystal structure of SARS-CoV-2 papain-like protease (PLpro) | Viral protein/protease |

The results of the molecular docking, the energy characteristics of the binding affinity in kcal/mol (A), and the binding affinity score (BAS) of the studied molecules for the main predicted categories of biological targets are presented in **Table 5**.

**Table 5.** Results of molecular docking: Energy characteristics of binding affinity in kcal/mol (A) and binding affinity score (BAS) of the studied molecules for the main predicted categories of biological targets.

| Biological targets |  | Ligand | Hit2 | Hit3 | Hit5 | Hit7 | Hit9 | Hit10 | Hit13 | Hit15 | CEP |
| --- | --- | --- | --- | --- | --- | --- | --- | --- | --- | --- | --- |
| 4E4L | A | -11.0 | -5.8 | -6.9 | -6.8 | -8.4 | -6.5 | -6.4 | -8.3 | -6.3 | -5.9 |
|  | BAS | -18.4 | -14.1 | -13.0 | -13.6 | -21.7 | -29.0 | -28.4 | -13.8 | -16.4 | -10.7 |
| 4D1S | A | -10.7 | -8.4 | -9.6 | -9.3 | -9.5 | -9.5 | -9.5 | -8.5 | -9.4 | -5.5 |
|  | BAS | -13.7 | -14.6 | -14.3 | -11.0 | -14.4 | -17.6 | -18.9 | -6.2 | -2.3 | -10.6 |
| 4Z16 | A | -8.2 | -8.7 | -9.5 | -9.0 | -6.9 | -9.8 | -9.4 | -9.3 | -9.2 | -10.4 |
|  | BAS | -10.9 | -18.3 | -12.1 | -14.7 | -12.4 | -13.9 | -12.5 | -13.3 | -10.2 | -10.6 |
| 1ALU | A | -4.6 | -5.1 | -5.9 | -6.1 | -6.0 | -5.8 | -6.1 | -6.9 | -5.7 | -4.5 |
|  | BAS | -12,5 | -2.1 | -16.8 | -7.6 | -15.3 | -2.1 | -3.2 | -7.9 | -1.0 | -2.0 |
| **7WOF** | **A** | -6.8 | -6.4 | -6.3 | -6.7 | -7.1 | -7.6 | -7.6 | -7.7 | -7.9 | -4.9 |
|  | **BAS** | -14.6 | -10.5 | -8.8 | -13.5 | -6.5 | -8.8 | -5.4 | -1.4 | -13.6 | 9.3 |
| **7LLZ** | **A** | -8.9 | -7.8 | -8.1 | -8.5 | -8.8 | -8.0 | -8.0 | -8.4 | -7.9 | -7.3 |
|  | **BAS** | -20.0 | -12.5 | -1.1 | -12.5 | -16.7 | -3.9 | -7.3 | -7.4 | -13.2 | -15.2 |

The tested molecules showed potential inhibition of Target 4Z16 (Jak3), a domain of the Janus kinase JAK3, a representative of tyrosine kinases, cytoplasmic enzymes involved in mediating intracellular signaling from cytokine receptors, which are responsible for inflammation processes [36]. Hit molecules also showed potential inhibition of SARS-COV-2 proteases Mpro and PLpro.

Several ADME parameters were calculated to evaluate the suitability of the identified compounds in terms of drug-like properties, including chemical absorption, distribution, metabolism, and excretion, which play a crucial role in determining potential drug molecules. **Table 6** presents parameters such as lipophilicity (clogP), topological polar surface area (TPSA), molecular weight (parameter SIZE, g/mol), solubility (LogS), fraction of sp3 carbon atoms (Fraction Csp3), flexibility, and optimal values of these parameters. The studied molecules conform to the drug-likeness properties.

**Table 6.** Drug-likeness properties of selected hits.

| **Name** | **Log P<sub>ow</sub>** | **SIZE (g/mol)** | **TPSA, Å<sup>2</sup>** | **LogS** | **Fraction C (sp<sup>3</sup>)** | **Flexibility** |
| --- | --- | --- | --- | --- | --- | --- |
|  | *-0.7 < X < 5.0* | *150 < X < 500* | *20 < X < 130* | *0 < -X < 6* | *0.25 < X < 1* | *0 < X < 9* |
| **Hit2** | 3.51 | 504.60 | 121.65 | -4.82 | 0.26 | 12 |
| **Hit3** | 5.09 | 442.51 | 90.02 | -5.71 | 0.23 | 9 |
| **Hit5** | 4.35 | 462.93 | 90.02 | -6.00 | 0.20 | 9 |
| **Hit7** | 5.00 | 389.47 | 97.34 | -5.72 | 0.14 | 5 |
| **Hit9** | 3.74 | 438.48 | 137.16 | -4.98 | 0.14 | 7 |
| **Hit10** | 3.61 | 450.51 | 116.39 | -4.89 | 0.18 | 8 |
| **Hit13** | 1.97 | 435.41 | 122.28 | -3.70 | 0.09 | 9 |
| **Hit15** | 3.22 | 473.57 | 151.90 | -4.63 | 0.25 | 9 |

### In vitro models

#### SARS-CoV-2 spike/ACE2 pseudovirus neutralization assay

The S-protein and ACE2 receptor play an essential role in the early phases of coronavirus infection [37]. The established binding assay is based on measuring luciferase activity and utilizes stable hACE-2 overexpressed HEK293T cells and SARS-CoV-2 S-protein expressing VSV-G pseudotyped lentiviruses. Omicron B1.1.529 SARS-CoV-2 coronavirus subtype was chosen as the most related subtype to the current variant. It is well known that the mutations increase the infectivity of the COVID-19 virus [38]. The cytotoxicity against hACE2-293T cells was tested, and the results revealed no toxicity (**Table 7**).

**Table 7.**
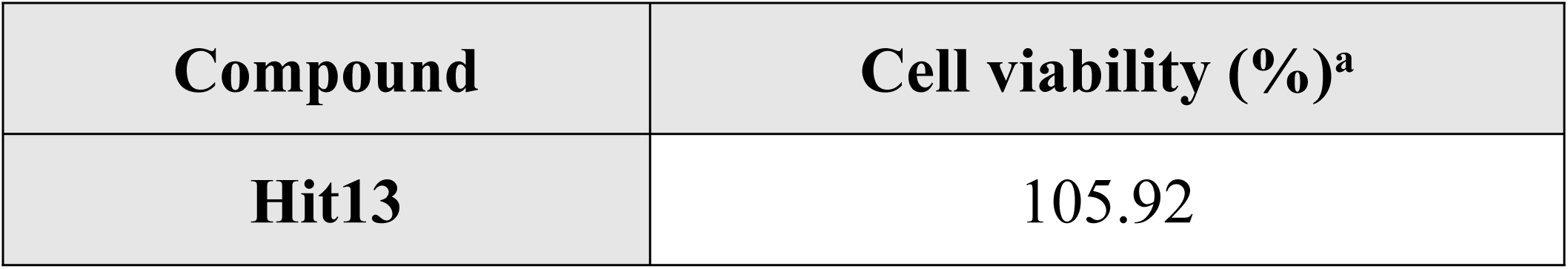
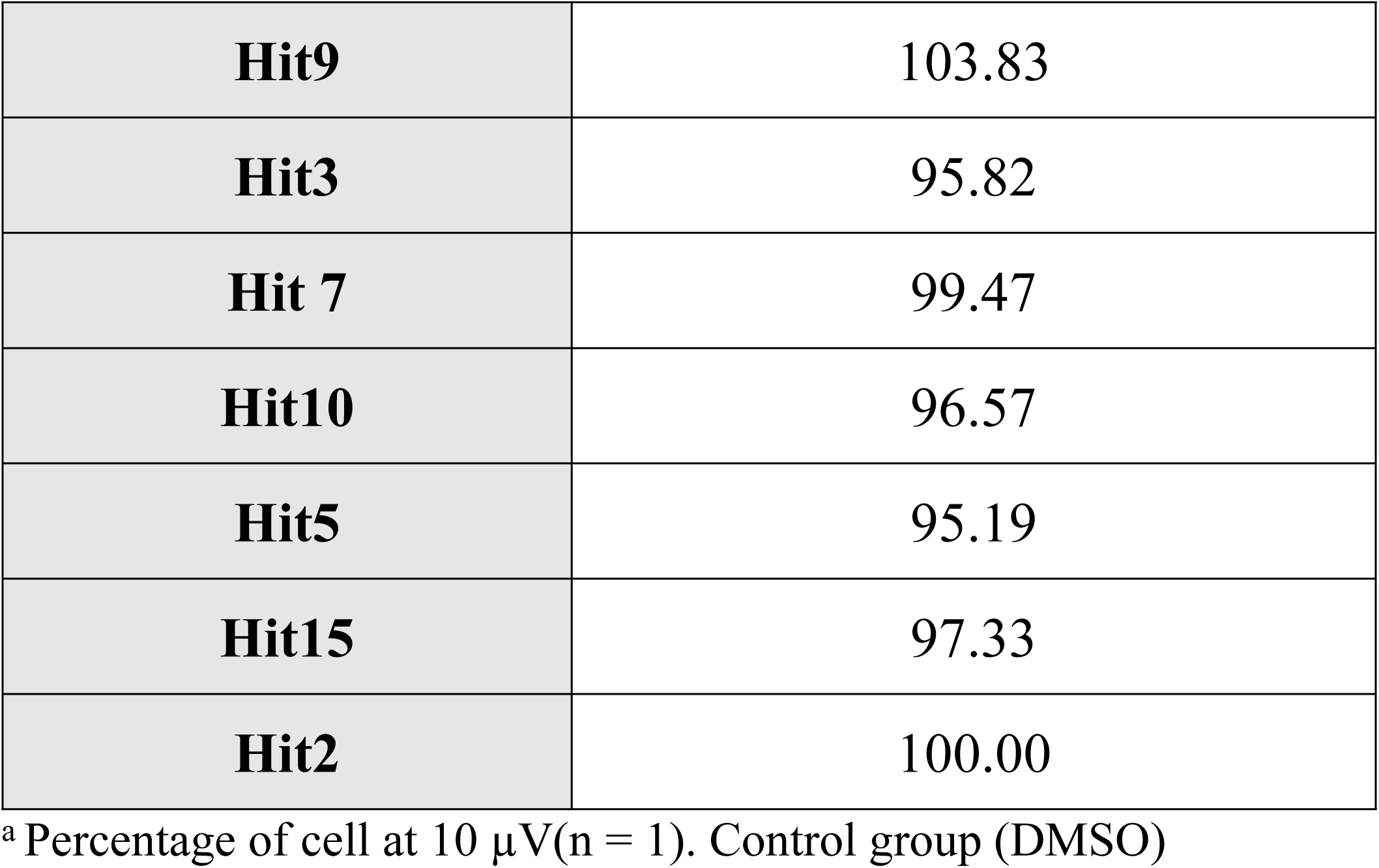
Effects of synthetic compounds on hACE-2 overexpressed HEK293T viability.

| **Compound** | **Cell viability (%)<sup>a</sup>** |
| --- | --- |
| **Hit13** | 105.92 |
| **Hit9** | 103.83 |
| **Hit3** | 95.82 |
| **Hit 7** | 99.47 |
| **Hit10** | 96.57 |
| **Hit5** | 95.19 |
| **Hit15** | 97.33 |
| **Hit2** | 100.00 |

<sup>a</sup> Percentage of cell at 10 $\mu$ V(n = 1). Control group (DMSO)

Next, the SARS-CoV-2 S-protein and ACE2 binding assay results showed activity of **Hit15** with 43.0% inhibition at 10 *μ*M (**Table 8**). The compound possesses sulfonyl and pyrimidone moiety that may be responsible for the observed activity [39, 40].

**Table 8.** Effect of synthetic compounds in the pseudovirus neutralization assay of Omicron variants (SARS-CoV-2 Spike protein pseudotyped lentivirus type).

| **Compound** | **Inh%<sup>a</sup>** |
| --- | --- |
| **Hit13** | 19.78 ± 8.29 |
| **Hit9** | 2.14 ± 10.15 |
| **Hit3** | 10.46 ± 5.69 |
| **Hit 7** | 22.12 ± 14.37 |
| **Hit10** | 14.25 ± 9.76 |
| **Hit5** | 0.08 ± 6.23 |
| **Hit15** | 43.00 ± 0.80 |
| **Hit2** | 29.56 ± 13.34 |
| **Cepharanthine** | 97.33 ± 0.08 |

<sup>a</sup> Percentage of inhibition (Inh.%) at 10 μM. Results are presented as mean ± S.E.M. (n = 2). Control group (DMSO). SARS-CoV-2 pseudovirus strain: Omicron (B.1.1.529).

In the pseudovirus neutralization assay, cepharanthine showed 97.3% inhibition at 10 *μ*M and IC_50_ 1.07 ± 0.24 µM. The antiviral potential of HIT15 and cepharanthine were in agreement with the reported data [41]. They indicated that both compounds may suppress infection of SARS-CoV-2 pseudovirus in hACE2-overexpressed host cells and may interfere with the S-protein/ACE2 binding. Previous research suggested that reducing the activity of TMPRSS2 using plant secondary metabolites could help manage COVID-19. TMPRSS2 is crucial for the virus entry stage by priming the S-protein of SARS-CoV-2, which facilitates the fusion of viral and host cell membranes. In an *in vitro* molecular docking study, cepharanthine interacted well with the TMPRSS2 and SARS-CoV-2 S-protein [43]. These data bode well with the docking result for cepharantine (**Table 3**) in the SARS-CoV-2 S-protein/ACE2 binding pocket. The anti-inflammatory activity was studied *in vitro* in a model of inhibition of superoxide anion release and elastase in human neutrophils. The effect of new molecules on superoxide anion generation and elastase release in activated human neutrophils was studied (**Table 9**). Superoxide anion generation and elastase release were induced by FMLP/CB and measured respectively. All data are expressed as mean ± S.E.M. (n = 3–4). \**P* < 0.05, \*\**P* < 0.01, and \*\*\**P* < 0.001 compared with the control.

**Table 9.**
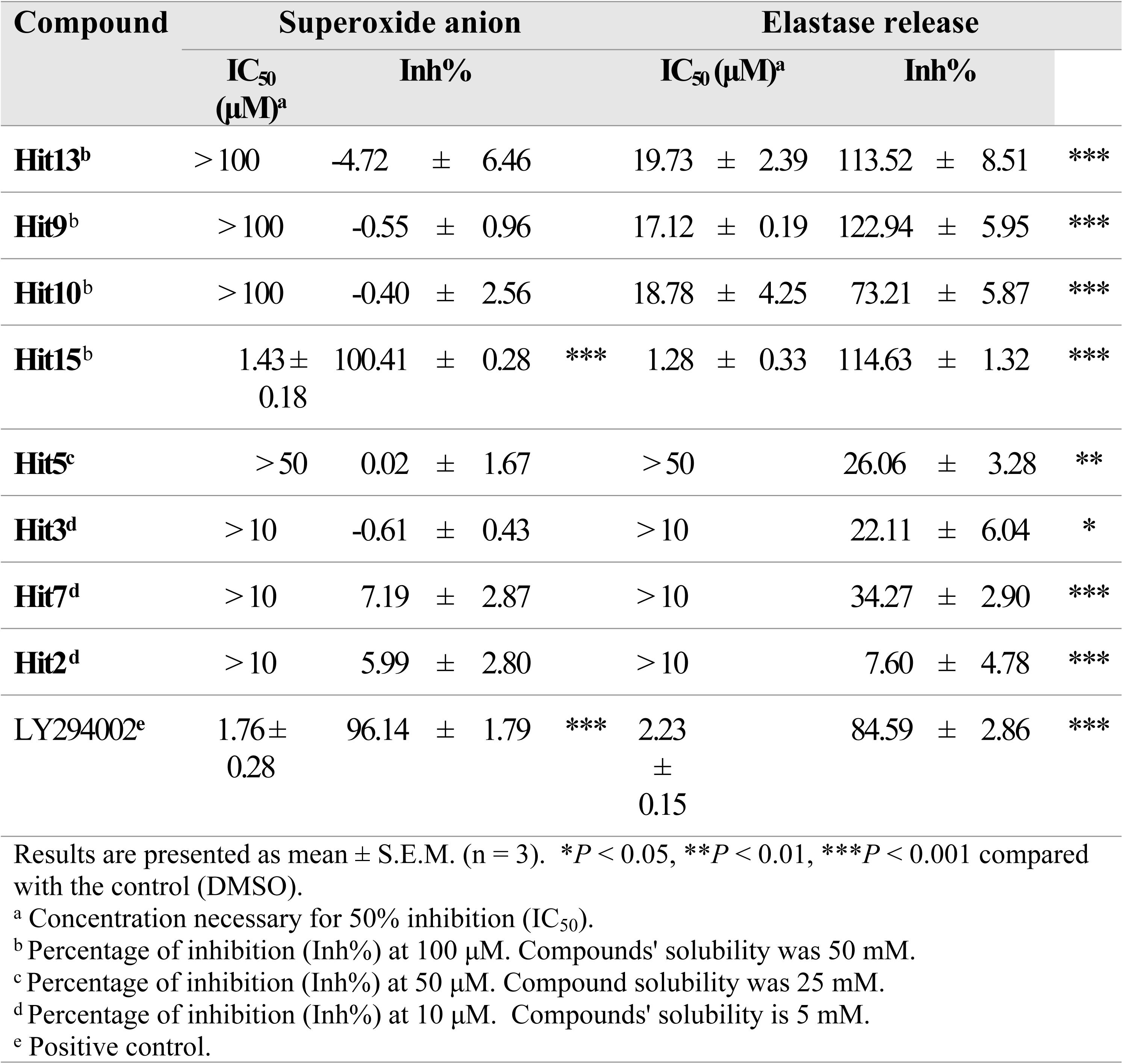
Effects of compounds on superoxide anion generation and elastase release in fMLF/CB-induced human neutrophils.

Experimental results showed that **Hit13**, **Hit9,** and **Hit10** moderately affected elastase release. **Hit15** showed good anti-inflammatory activity with IC_50_ 1.76 and 2.23 μM in both superoxide anion generation and elastase release assays in fMLF/CB-induced human neutrophils, respectively. The **Hit15** molecule has good potential for further development to treat COVID-19.

## CONCLUSION

A virtual *in silico* screening and receptor-oriented docking was conducted on three-dimensional structural models of active sites in biological molecules involved in the mechanisms of SARS-CoV-2 impact on the body, using cepharanthine as a representative structure. Several hit molecules were identified, characterized by binding energies with the active sites of the targets at levels comparable to or exceeding those of the representative cepharanthine structure. Receptor-oriented docking revealed that these hit molecules have the potential to inhibit SARS-CoV-2 proteases Mpro and PLpro, which play a crucial role in viral replication mechanisms, as well as to inhibit Janus kinase (Jak3), which mediates intracellular signal transduction and is responsible for inflammatory processes.

The hit molecules were evaluated *in vitro* for anti-inflammatory activity. They were demonstrated by assessing their inhibitory effects on elastase release in human neutrophils and the generation of superoxide anions, a characteristic of anti-inflammatory action. It was shown that the 2-((5-((4-isopropylphenyl)sulfonyl)-6-oxo-1,6-dihydropyrimidin-2-yl)thio)-N-(3-methoxyphenyl)acetamide (**Hit15**) exhibited potent anti-inflammatory activity. Several other molecules also demonstrated moderate effects on elastase release.

## Author Contributions

V.A.G., V.V.I. - Conceptualization,

Y.-L.C., S.-Y.F., L.V.Y., O.M.K. - Data Curation

V.A.G, O.M.K. - Project Administration

V.V.I., A.B.Z., D.O.A. - Software

O.M.K., T.-L.H. - Supervision.

L.V.Y., V.V.I., S.M.K., M.E.-S., M.K., O.O.M. - Writing – original draft

V.A.G. – Writing - Review & Editing;

T.-L.H., O.M.K. - Funding Acquisition

All authors have read and agreed to the published version of the manuscript.

## Acknowledgement

VVI, ABZ, SMK, L.V.Y and O.M.K. express their gratitude to the National Research Foundation of Ukraine (https://nrfu.org.ua/en/fundraising_en/) for financial support (grant No. 87/0062 (2021.01/0062) “Molecular design, synthesis and screening of new potential antiviral pharmaceutical ingredients for the treatment of infectious diseases COVID-19”). This research was supported by grants from the National Science and Technology Council (https://www.nstc.gov.tw) (NSTC 113-2321-B-255-001, 113-2321-B-182-003, 112-2321-B-182-003, 112-2321-B-255-001, 111-2320-B-255-006-MY3, and 111-2321-B-255-001 granted to T.L.H.; and 113-2320-B-037-023, 112-2320-B-037-012, and 111-2320-B-037-007 granted to M.K.), Chang Gung University of Science and Technology (https://english.cgust.edu.tw) (ZRRPF3L0091 and ZRRPF3N0101) granted to T.L.H., Chang Gung Memorial Hospital (https://www.cgmh.org.tw/eng) (CMRPF1P0051-3, CMRPF1P0071-3) granted to T.L.H., and Kaohsiung Medical University Research Foundation (https://www.kmu.edu.tw/index.php/en-gb/research) (KMU-Q113011) granted to M.K., Taiwan. We thank Prof. T. Langer for the opportunity to work with the LigandScout suite.

The funders had no role in the study design, data collection, analyses, decision to publish, or manuscript preparation.

## Institutional Review Board Statement

The study was conducted according to the guidelines of the Declaration of Helsinki and approved by the Institutional Review Board of Chang Gung Memorial Hospital (Registration number: IRB No.: 202301906A3C501), 1^st^ August to 30^th^ September 2024 was the recruitment period for this study.

## Informed Consent Statement

Informed consent written and signed by donors was obtained from all subjects involved in the study. No minors were included.

## Data Availability Statement

All data are fully available without restriction. All relevant data are within the manuscript.

